# Expression profile analysis of COVID-19 patients

**DOI:** 10.1101/2023.12.03.23299339

**Authors:** Valentin S. Shimansky, Oleg S. Popov, Tatiana G. Klochkova, Svetlana V. Apalko, Natalya N. Sushentseva, Anna Yu. Anisenkova, Sergey V. Mosenko, Sergey G. Shcherbak

## Abstract

The course of COVID-19 is determined by various factors. Studies worldwide have shown a correlation between changes in the expression profile and the severity of the disease. Thus, an in-depth study of differentially expressed genes will allow a more detailed investigation of the metabolic changes occurring in the background of coronavirus infection. The technique of RNA sequencing and subsequent bioinformatics analysis is suitable for such research tasks. In our study, we compared groups of samples from patients with mild and severe disease course and identified a number of differentially expressed genes. These genes are involved in metabolic pathways responsible for immune response, signaling, intercellular communication, metabolism of various compounds etc. We then identified master regulators whose function and role in pathways enriched by differentially expressed genes makes them potential targets for biochemical and meta-studies.

## INTRODUCTION

Work on the peripheral transcriptomic signatures of COVID-19 is being conducted by many research groups. Many different aspects of the disease are being investigated. For example, in a study by Chinese scientists on the peculiarities of protein ubiquitination in COVID-19, comparing gene expression in peripheral blood mononuclear cells of 4 patients with severe disease and 4 healthy controls, 268 differentially expressed genes were identified. As a result, we were able to identify 6 transcription factors and 2 microRNAs that are key factors in the regulation of ubiquitination in patients with severe COVID-19^1^. A study of differentially expressed matrix RNAs, microRNAs and long non-coding RNAs by researchers from China identified 25,482 mRNAs, 23 microRNAs and 410 long non-coding RNAs whose expression levels differed between COVID-19 patients and control donors. mRNAs that are overexpressed in samples of COVID-19 patients are mainly involved in antigen processing and endogenous antigen presentation, positive regulation of T-cell-mediated cytotoxicity, and positive regulation of gamma delta T-cell activation. mRNAs with reduced expression are mainly involved in glycogen biosynthesis ^2^.

A study of the blood transcriptome of people of different ages in the context of measuring the expression level of genes whose products interact with SARS-CoV-2 viral structures revealed five genes that change their expression with aging. They are involved in immune response, inflammation, cellular component and cell adhesion, and platelet activation/aggregation. Moreover, the expression profile changed differently in men and women with aging ^3^. In general, the transcriptomic signature of COVID-19 overlaps to a large extent with that of influenza and other acute respiratory infections. Studies of dynamic changes in transcriptome of COVID-19 patients are beginning to appear. The data from these studies show that the blood transcriptome undergo dramatic and consistent changes as in the early stage of recovery. And even months after clinical recovery, gene expression levels does not return to the values of a healthy person ^4^. The development of interpretable transcriptome panels for profiling the immune response to SARS-CoV-2 infection is already underway, with target genes categorized into groups: immunologic relevance, role in disease progression, and interaction with SARS structures ^5^.

Transcriptomic analysis can be a powerful approach to assess the molecular response of the host and can provide valuable information both on the pathophysiology of COVID-19 and find master regulators that could be potential targets for effective therapies. The aim of this study was to analyse differential gene expression to identify a set of regulatory genes affecting key molecular and cellular pathways involved in the pathogenesis of COVID-19.

All patients signed informed voluntary consent. The study was approved by the expert ethics board of the St. Petersburg State Health Care Institution “City Hospital No. 40” (protocol No. 171 dated May 18, 2020). Biomaterial from the biobank collection in St. Petersburg State Health Care Institution “City Hospital No. 40” was used to fulfil the study objectives.

## RESULTS

### Participant Characteristics

The participants of the study were the patients of the infectious disease department of the St. Petersburg State Health Care Institution City Hospital No. 40, Kurortny District who were admitted for treatment with coronavirus infection (confirmed by polymerase chain reaction). The studied patients were divided into two groups according to the severity of the disease (group 1 - mild and moderate course, group 2 - severe and extremely severe course of the disease). Age-sex structure of the groups is presented in Table 1.

**Table 1:**
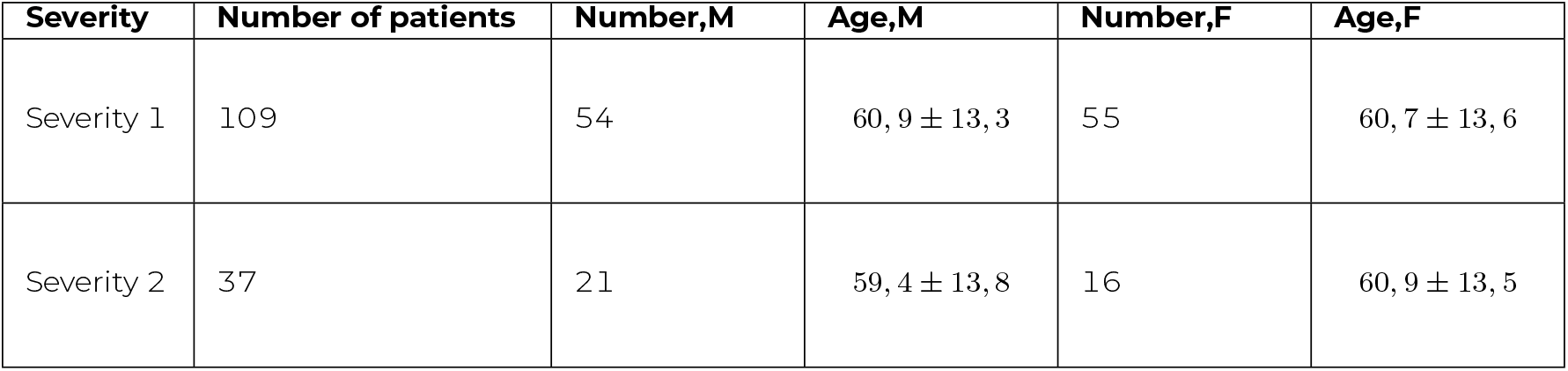
Age-sex structure of the groups.

### Results of differential expression analysis

We obtained the following results analysing blood samples from patietns divided by severity. 4734 genes passed the 0.01 statistical significance level. 806 genes were upregulated and 3925 genes were downregulated in group 2 relative to group 1. Upregulated genes with the lowest p-value are listed in Table 2. Downregulated genes with the lowest p-value are listed in Table 3.

**Table 2:** Top 10 upregulated genes by p-value.

| Gene name | p-value | logFC |
| --- | --- | --- |
| *ALOX15* | 2.57e-07 | 1.314 |
| *SLC4A10* | 4.96e-07 | 0.829 |
| *PID1* | 9.82e-07 | 0.516 |
| *TRAV1-1* | 1.24e-06 | 0.455 |
| *SIGLEC8* | 1.35e-06 | 1.33 |
| *TRBV29-1* | 1.49e-06 | 0.315 |
| *GIMAP5* | 1.67e-06 | 0.047 |
| *FCER1A* | 1.81e-06 | 0.538 |
| *CCR3* | 2.00e-06 | 0.690 |
| *SMPD3* | 2.55e-06 | 0.461 |

**Table 3:** Top 10 downregulated genes by p-value.

| Gene name | p-value | logFC |
| --- | --- | --- |
| *TRBJ1-6* | 2.12e-06 | -1.17e-01 |
| *NR3C2* | 2.91e-06 | -2.33e-02 |
| *PLXDC1* | 3.35e-06 | -1.74e-01 |
| *TRBJ1-3* | 5.98e-06 | -3.57e-03 |
| *ADAMTS2* | 7.67e-06 | -5.69e-01 |
| *LINC00649* | 8.17e-06 | -4.90e-01 |
| *TRAJ10* | 8.38e-06 | -7.85e-02 |
| *RORC* | 8.48e-06 | -5.51e-01 |
| *LINC01550* | 9.74e-06 | -2.88e-01 |
| *BACH2* | 9.96e-06 | -5.78e-01 |

GO enrichment analysis showed that upregulated genes are grouped in pathways responsible for adaptive immune response, signalling, intercellular communication, and several others (Table 4).

**Table 4:** Top GO terms, enriched with upregulated genes.

| Metabolic pathway | Number of hits | GO group size | p-value |
| --- | --- | --- | --- |
| Regulation of cell migration | 39 | 922 | 3.14e-05 |
| Chemotaxis | 28 | 707 | 0.001 |
| Cell part morphogenesis | 29 | 724 | 0.001 |
| cellular nitrogen compound metabolic process | 11 | 150 | 0.002 |
| Regulation of MAPK cascade | 30 | 782 | 2.04e-05 |
| Regulation of development process | 76 | 2 860 | 1.68e-05 |
| Response to growth factor | 28 | 725 | 0.002 |
| Ventral trunk neural crest cell migration | 3 | 4 | 0.001 |
| Regulation of signaling | 96 | 3 927 | 0.002 |
| Trigeminal nerve structural organization | 3 | 5 | 4.47e-05 |
| Adaptive immune response | 50 | 978 | 3.14e-09 |
| Trigeminal nerve morphogenesis | 4 | 12 | 3.37e-05 |
| Immune response | 81 | 2 893 | 1.91e-04 |
| Signaling | 180 | 7 537 | 5.14e-07 |
| Cell communication | 179 | 7 554 | 8.51e-07 |
| Signal transduction | 167 | 6 942 | 2.58e-09 |
| Response to stimulus | 226 | 10 504 | 1.40e-06 |
| Cell surface receptor signaling pathway | 102 | 3 548 | 4.53e-09 |

Downregulated genes are concentrated in molecular pathways responsible for the processes of cellular nitrogen and heterocycle compound metabolism, gene expression, etc. (Table 5).

**Table 5:**
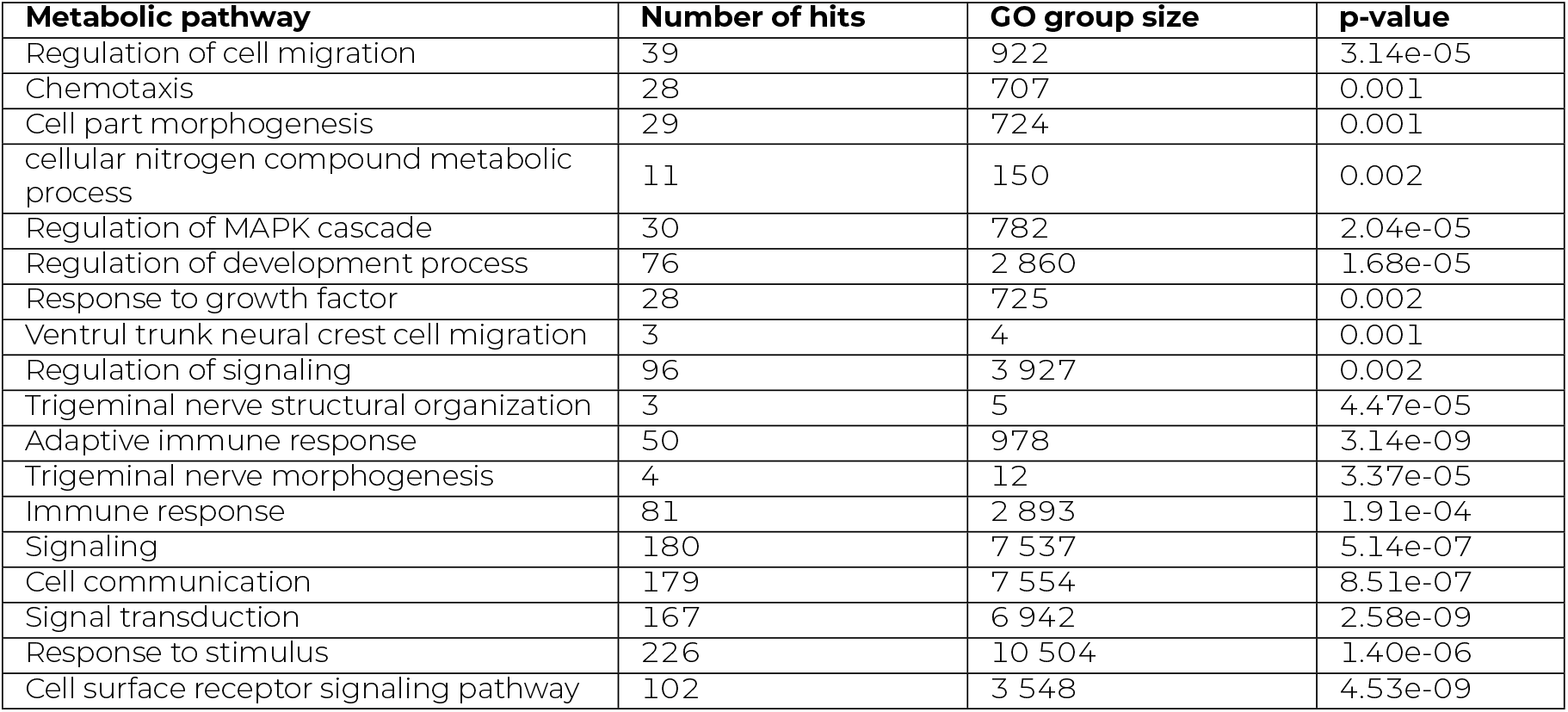
Top GO terms, enriched with downregulated genes.

| Metabolic pathway | Number of hits | GO group size | p-value |
| --- | --- | --- | --- |
| Regulation of cell migration | 39 | 922 | 3.14e-05 |
| Chemotaxis | 28 | 707 | 0.001 |
| Cell part morphogenesis | 29 | 724 | 0.001 |
| cellular nitrogen compound metabolic process | 11 | 150 | 0.002 |
| Regulation of MAPK cascade | 30 | 782 | 2.04e-05 |
| Regulation of development process | 76 | 2 860 | 1.68e-05 |
| Response to growth factor | 28 | 725 | 0.002 |
| Ventrul trunk neural crest cell migration | 3 | 4 | 0.001 |
| Regulation of signaling | 96 | 3 927 | 0.002 |
| Trigeminal nerve structural organization | 3 | 5 | 4.47e-05 |
| Adaptive immune response | 50 | 978 | 3.14e-09 |
| Trigeminal nerve morphogenesis | 4 | 12 | 3.37e-05 |
| Immune response | 81 | 2 893 | 1.91e-04 |
| Signaling | 180 | 7 537 | 5.14e-07 |
| Cell communication | 179 | 7 554 | 8.51e-07 |
| Signal transduction | 167 | 6 942 | 2.58e-09 |
| Response to stimulus | 226 | 10 504 | 1.40e-06 |
| Cell surface receptor signaling pathway | 102 | 3 548 | 4.53e-09 |

Master regulator analysis showed the following results: *CDH2, CXCL9, KIT, IL13, EPHA4, FLT3, DUSP6, FGFR2, IL4* for upregulated genes; *RHOH, NFATC3, FXN, ADRB2, NTRK1, NAA30, ZC3H12D, MAF, KLRD1* for downregulated genes.

## MATERIALS AND METHODS

### RNA extraction and sequencing

RNA was isolated manually from blood samples using the phenol-chloroform extraction method. Library preparation was performed using KAPA RiboErase HMR reagents (Roche), RNA Hyper Prep (Roche), KAPA Universal Adapter (Roche), KAPA UDI Primer Mixes (Roche). Conversion was performed using High-Throughput Sequencing Primer Kit (App-C) and MGIEasy Universal Library Conversion Kit (App-A) (MGI) reagents. Sequencing of the resulting libraries was performed on an MGISEQ-2000 sequencer using a 100 base pair-end length read on a DNBSEQ-G400 High-throughput Sequencing Set (PE100, 360 GB) cell (MGI).

### Bioinformatics analysis

Quality control was performed for raw reads using the FastQC program. Alignment to the human genome hg38 version was performed using the STAR aligner with standard settings. Counting reads and gene annotation were performed with the featureCounts program.

### Statistical analysis

The nonparametric Mann-Whitney test was used to calculate statistically significant differences in gene expression, and the log fold change (logFC) was used to measure the differential expression. Calculations were performed in the R software package. GO pathway enrichment analysis and the master regulator analysis were performed on the ge.genexplain.com platform.

## Data Availability

All data produced in the present study are available upon reasonable request to the authors.

## DATA AND CODE AVAILABILITY

Personal genetic and clinical data are under restrictions and are available through collaboration with the St. Petersburg State Health Care Institution City Hospital No. 40, Kurortny District hospital.

## CONFLICT OF INTEREST

The authors declare that they have no competing interests.

## ACKNOWLEDGMENTS

The authors are grateful to the study participants and the staff from the St. Petersburg State Health Care Institution City Hospital No. 40, Kurortny District hospital. The authors would like to thank all authors of the included studies for their valuable contributions to data collection. This work was supported by Saint Petersburg State University, project ID: 94029859.

## Notes

### Competing Interest Statement

The authors have declared no competing interest.

### Author Declarations

The study was approved by the expert ethics board of the St. Petersburg State Health Care Institution "City Hospital No. 40" (protocol No. 171 dated May 18, 2020).

### Summary of Updates

Added affilations and acknowledgements

